# Variance Estimation for Assessing Healthcare Providers’ Performance using log Standardized Incidence Ratio

**DOI:** 10.1101/2025.04.22.25326183

**Authors:** Solomon Woldeyohannes, Yomei Jones, Paul Lawton

## Abstract

**Introduction:** In healthcare providers’ performance assessment, standardized incidence ratios (SIRs) are essential tools used to assess whether observed event rates deviate from expected values. Accurate estimation of variance in these ratios is crucial as it affects decision-making regarding providers’ performance. There is little data on how the choice of these variance estimation methods affects decision-making. In this paper, we compared three methods, namely, delta-method, bootstrapping and Bayesian approaches, to estimate the variance of the logarithm of SIR (Log-SIR).

**Methods and analysis:** Using patient-level data from Australia and New Zealand Dialysis and Transplant Registry (ANZDATA) for 2005-2023, we used a random effects model to predict treatment at home one year after starting treatment. We compared the three approaches (with over 5000 iterations for bootstrapping and MCMC sampling) using bias, variance and mean square errors (MSE) as performance measures. Using the three methods, funnel plots were used to compare the hospitals’ performance in treating Indigenous and non-Indigenous patients close to home, as a service-level measure of equity.

**Results:** The bias values across all methods are similar, with Bayesian narrowly having the lowest bias (0.01922), followed by the delta-method (0.01927) and Bootstrap (0.02567). In addition, the Bayesian exhibits the lowest variance (0.00005), indicating more stable and less dispersed estimates. The delta-method has a higher variance (0.00016), while Bootstrap has the highest variance (0.00027), meaning it introduces more uncertainty. Finally, the Bayesian has the lowest MSE (0.00042), indicating better overall accuracy while the Bootstrap has the highest MSE (0.00094), showing it is the least reliable method.

**Conclusion:** We demonstrate that these methods can be used to measure equity for patient-centred outcomes, both within and between service providers simultaneously. The choice of variance estimation method is critical and heavily affects the interpretation of the performance of health service providers. We favour the Bayesian MCMC method. The Bayesian MCMC method found to be better approach.

## 1 INTRODUCTION

Public scrutiny of healthcare service performance have been emphasized in the last two decades. For instance, the Australian government has introduced the National Health Reform in 2011^1^ and recently the 2020–25 National Health Reform Agreement (NHRA) ^2^. This in turn has led to increased attention to institutional comparisons on the basis of quantitative outcome measures such as standardized mortality ratios (SMRs) in which, with the aid of confidence intervals, ‘outlying’ institutions are identified ^3^. A league table of hospitals based on mortality ^4^ and Shewhart’s control charts (using 1, 2 & 3 SD limits) ^5^ have been proposed and criticised to compare institutional ranking. More recently, a ‘funnel plot’, in which an estimate of an underlying quantity is plotted against an interpretable measure of its precision has become a useful graphical aid for institutional comparisons ^3,6,7^. Although, funnel plots have been used in meta-analyses, in particular to detect publication bias, they have recently been strongly recommended as the most appropriate way to display performance indicators such as comparisons of risk-adjusted rates between healthcare units ^8^. SMRs are the commonly used performance index for institutional comparisons ^9^. However, this concept has been readily extended to encompass several other indices such as age-standardised relative survival and excess hazard ratios (EHRs) ^8^, and standardised incidence ratio (SIR) ^10^. Estimating the variance of log SIR (denotated by Log-SIR hearafter) is necessary for creating false discovery rates (FDRs) in studies that use funnel plots for assessing centres’/hospitals’ performance. Accurate estimation of variance in these ratios is crucial as it affects decision-making regarding hospital performance and quality improvement strategies. Despite different variance estimation methods being used widely in application, there are no data on how the choice of these methods affects the assessment of performance. In this paper, we compared three methods, namely, delta-method, bootstrapping and Bayesian approaches, to estimate the variance of the Log-SIR and subsequent funnel plot approaches to build false discovery rates for the Log-SIR.

The delta-method is the analytical approache to estimate the variance of the logarithm of SMR, denoted by Log-SMR hereafter. It approximates the variance of a function of random variables by using the Jacobian matrix and the covariance matrix of the original variables ^11^.

Quaresma, et al. ^12^ used the delta-method directly to estimate risk-adjusted excess hazard ratios (EHRs) as performance measure in a study of population-based cancer survival. Also, Powell et al. ^13^ applied the delta-method to approximate the variance of demographic parameters in avian biology studies. Vasilevskis, et al. ^9^ used confidence intervals (CIs) for comparing SMR using bootstrapping approach in a study involving prediction of 30-Day ICU Mortality. Though confidence intervals can be constructed for SMR directly, Hosmer and Lemeshow demonstrated CIs with good coverage for the logarithm of the SMR ^14^. Also, Austin ^15^ investigated four bootstrap procedures for estimating confidence intervals for predicted-to-expected ratios in a hospital profiling study. He indicated that existing bootstrap procedures should not be used to compute confidence intervals for predicted-to-expected ratios when conducting provider profiling ^15^.

Similar to bootstrapping, a Bayesian approach via MCMC can be used to approximate this variance. For instance, Massimo, et al. (2011) applied Bayesian hierarchical models to estimate small area level SMR and constructed False Discovery Rates (FDRs) in a study of liver cancer morbidity cases recorded between 1998 and 2003 in Emilia–Romagna municipalities ^16^. In addition, Suku, et al. ^17^ demonstrated the application of a Bayesian hierarchical model in assessing hospital and operator variation in cardiac rehabilitation referral and participation after percutaneous coronary intervention (PCI) using a retrospective observational cohort of patients who underwent PCI at 48 non-federal Michigan hospitals between January 1, 2012 and March 31, 2018.

Applying the delta-method depends on fulfillment of underlying distributional assumption; asymptotic normality. Bootstrapping, on the other hand, has the advantage of not relying on distributional assumptions and can be used to directly estimate the distribution of Log-SIR or Log-SMR. This can lead to more robust variance estimates, particularly in settings with small sample sizes or unknown distributions. By resampling, bootstrapping accounts for sampling variability and can help improve the precision of performance assessments ^11^. Therefore, this paper compares the three variance estimation methods using bias, variance and mean square error (MSE) as measures of performance.

For over 25 years, First Nations health organisations and patients in rural and remote Australia have persistently called for more responsive treatment, closer to home, for First Nations peoples with end-stage kidney disease ^18,19^. Community-led advocacy groups have continued this call in more recent years. A national meeting of First Nations kidney failure patients in September 2017 renewed this message ^20^. Over the last 15 years, substantial progress has been made in expanding and decentralising haemodialysis care across remote Australia ^21^. Nevertheless, most treatment is still provided as haemodialysis in nurse-facilitated centres in major or regional towns, rather than at home in remote communities ^22^.

The Return to Country Study, of which this methodological work is a part, aims to characterise the socioeconomic, environmental, health service and biomedical factors driving the health outcomes and patterns of health service utilisation experienced by First Nations Australians receiving kidney replacement therapy (KRT) and investigate whether health service changes to address these identified barriers can achieve higher rates of KRT closer to home ^23^.

This study is funded by the National Health and Medical Research Council of Australia (GNT1158075). Ethics approval has been obtained so far from the Human Research Ethics Committee (HREC) of the Northern Territory Department of Health and Menzies School of Health Research (2019-3530), Far North Queensland HREC (2023/QCH/99606 (Nov ver 4)-1732), the Central Adelaide Local Health Network HREC (2023/HRE00209), the Aboriginal Health Council of South Australia (AHREC Protocol #: 04-23-1078), the Aboriginal Health and Medical Research Council of New South Wales (AH&MRC HREC reference: 2230/24) and the Far North Queensland Human Research Ethics Committee (FNQ HREC reference: HREC/2023/QCH/99606 (Nov ver 4)-1732).

Trial registration number ACTRN12623001241628.

## 2 METHODS

### 2.1 Data source

The source of data for our motivating example is the ANZDATA registry ^6,23^. The ANZDATA Registry receives, collates and analyses data from centres providing care for patients receiving long-term dialysis or kidney transplantation in Australia and New Zealand. Data submission is voluntary but complete. For this methodological study, we used the data extract provided by ANZDATA for the Return To Country Study (ANZRREQ-471) ^23^.

In the following we presented model specification, the derivation of the variance for the Log-SIR using the delta-method, and a description of the bootstrap and Bayesian approaches for estimating variance of Log-SIR.

### 2.2 Model specification and Likelihood definition

Since we have binary outcome of receiving treatment close to home for ESKD, denoted by *y*_*ci*_, from *n*_*c*_ number of patients receiving treatment from centre *c*. Where, *c*, …, *N* centres, we proposed a Bernoulli sampling distribution for the probability of getting treatment close to home for the *i*^*th*^ patient from centre *c*. That is, *y*_*ci*_ ∼ Bernoulli(*p*_*ci*_) and a random effects logistic regression model can be specified as:

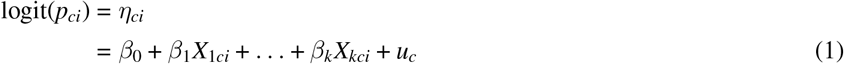

where: *y*_*ci*_ is the binary outcome for patient *i* in centre *c, X*_1*ci*_ … *X*_*kci*_ are *k* covariates for patient *i* in centre *c, β*_0_, *β*_1_, …, *β*_*k*_ are fixed effects, *u*_*c*_ is the random effect for centre *c*, assumed to be normally distributed: *u*_*c*_ ∼ *𝒩* (0, *σ*^2^) and *p*_*ci*_ = *P*(*y*_*ci*_ = 1).

Furter details of the model specification can be found in Appendix A.

Estimates of the fixed effects *β* and the variance parameter 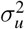 were obtained using maximum likelihood estimation (MLE) through the ‘glmer’ function in R ^24,25^.

Application of random intercept multilevel logistic regression models for indirectly standardise performance measures is explored in Clark, D.E., & Moore, L. ^26^ using National Trauma Data Bank data for admission year 2008. Zang et al. (2013) explored hierarchical logistic regression (LR) modelling under various conditions applying Bayesian and frequentist’s methods Yang, et al. ^27^

### 2.3 Delta-method for the variance of the Log-SIR)

The delta method is a technique used to approximate the variance of a function of one or more random variables ^28,29,30^. The first-order Taylor series approximation for moments of ratio estimators is used to derive the mean and variance estimates, see ^28^, pages 244-245. In the context of estimating the variance of the Log-SIR, we can apply the delta method to approximate the variance of log 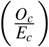.It approximates the variance of a function of random variables by using the Jacobian matrix and the covariance matrix of the original variables Boos ^31^, page 14. Accordingly, the variance of Log-SIR_*c*_ is approximated by:

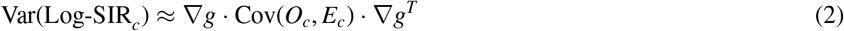

Where the covariance matrix of *O*_*c*_ and *E*_*c*_ is specified as:

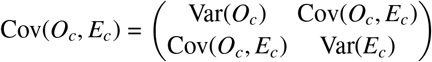

And the Jacobian matrix (gradient) ∇*g* of the function *g*(*O*_*c*_, *E*_*c*_) with respect to *O*_*c*_ and *E*_*c*_ is given by:

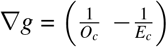

Substituting ∇*g* and Cov(*O*_*c*_, *E*_*c*_) into the formula, we get final expression for the variance:

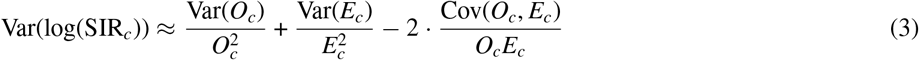

Detailed derivation of the final formula for the variance of log(SIR) using the delta method given the model specification and the likelihood formulations above is presented in Appendix B.

The next section summarizes the estimates for the Var(*O*_*c*_), Var(E_*c*_) and Cov(O_*c*_, E_*c*_).

#### 2.3.1 Variance of *O*_*c*_: Var(*O*_*c*_)

Let *Y*_*i*_ be the binary outcome for individual *i* in centre *c*. The observed incidence *O*_*c*_ is the sum of binary outcomes *Y*_*i*_ for individuals within the *c*^*th*^ centre. If patients share hospital-level characteristics, the outcomes *Y*_*i*_ are not independent but are correlated due to the shared random effect. The observed counts for centre *c* are:

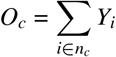

The variance of *O*_*c*_ is given by:

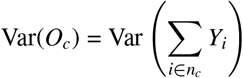

Using the property of variance for the sum of random variables, this expands to:

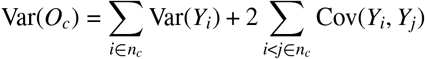

This expression is derived from the formula for the variance of the sum of random variables. Here, Var(*Y*_*i*_) represents the variance of the individual observations, and Cov(*Y*_*i*_, *Y*_*j*_) is the covariance between pairs of observations. The factor of 2 in front of the covariance term accounts for the fact that each covariance term is counted only once when summing over pairs *i* < *j*.

For a logistic regression model with random intercepts, the variance and covariance terms are:

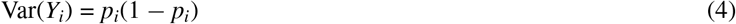

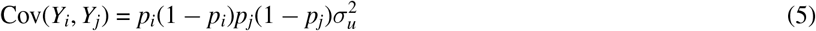

Thus,

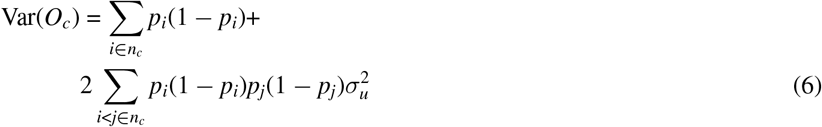

#### 2.3.2 Derivation of Var(*E*)

The expected counts *E* are the sum of predicted probabilities *p*_*i*_ for individuals within a centre. The variance of *E* arises from the uncertainty in the predicted probabilities due to the random effects.

The expected counts for centre *c* are:

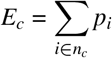

The variance of *E*_*c*_ is:

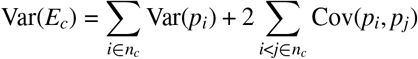

For the random-effects logistic regression model:

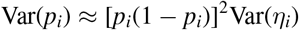

where 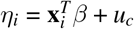 is the linear predictor. The covariance between *p*_*i*_ and *p*_*j*_ (for *i ≠ j*) is:

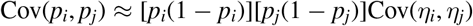

Since *η*_*i*_ and *η*_*j*_ share the same random effect *u*_*c*_:

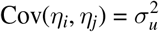

Thus:

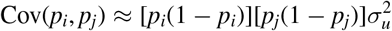

Combining these results:

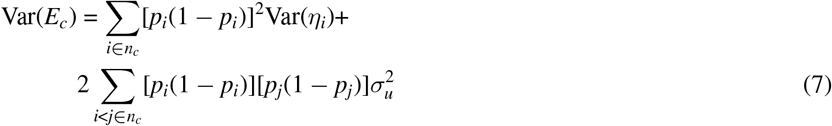

#### 2.3.3 Derivation of Cov(*O*_*c*_, *E*_*c*_)

The covariance between *O*_*c*_ and *E*_*c*_, where *O*_*c*_ is the observed count and *E*_*c*_ is the expected count for centre *c*, arises because both depend on the same underlying probabilities *p*_*i*_, wh are influenced by the shared random effect.

To derive the covariance Cov(*O*_*c*_, *E*_*c*_), given 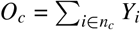 (Observed count) and 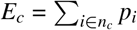 (Expected count), we have the covariance between *O*_*c*_ and *E*_*c*_ defined as:

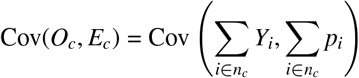

And using the property of covariance for sums, we get:

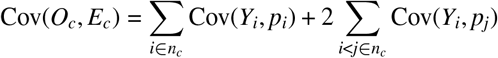

Therefore, the final expression of Cov(*O*_*c*_, *E*_*c*_) becomes :

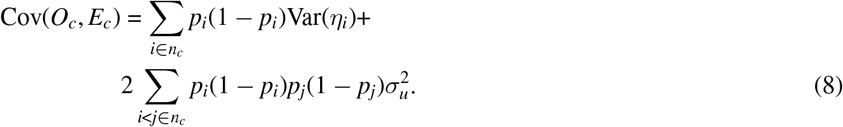

### 2.4 Bootstrapping approach

Commonly, bootstrap approach is used to approximate variance of the log standardized incidence ratio. By sampling with replacement from the observed sample, creating a resampled dataset of size *n* and repeating this *B* times, it creates a non-parametric bootstrapped distribution ^28^, pages 479-480. This distribution can be used to estimate the variance of the Log-SIR_*i*_. Mathematically, this can be summarised as:

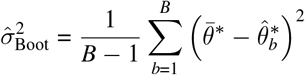

with 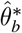 the Log-SIR_*c*_ estimate estimated in the *b*^*th*^ bootstrap sample and 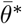 the mean Log-SIR_*c*_ estimate over the *B* bootstrap samples; here *B* = 5, 000.

### 2.5 Bayesian approach

Given the model specification given in (1), the posterior distribution for a random effects logistic regression model can be expressed in a hierarchical form, integrating over the random effects *u*_*c*_. It can be recalled that the form of a posterior for hierarchical models is ^31^:

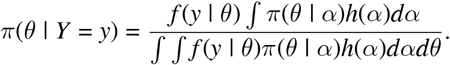

Using the likelihood for random effects logistic regression and priors for ***β*** and *u*_*c*_, the full posterior distribution can be shown to be:

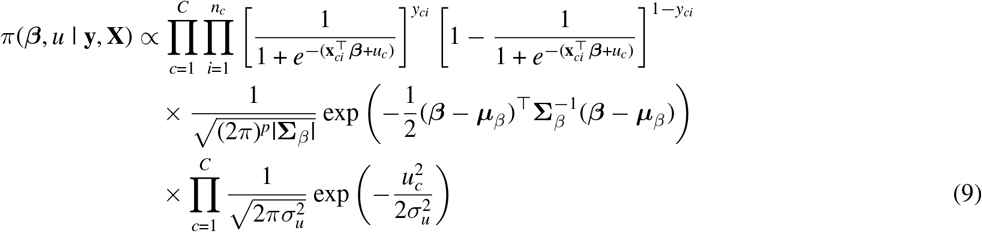

Details of the derivation of the full posterior distribution is summarized in supplementary material Appendix C.

Due to the need to integrate out the nuisance parameters in (9) and lack of conjugate priors, and the hierarchy involved, computing difficult integrals is required using Markov chain Monte Carlo (MCMC) methods whereby a dependent sequence of random variables are obtained with the property that in the limit these random variables have the posterior distribution.

Accordingly, the following information is used to estimate the variance of the Log-SIR using the Bayesian approach: *y*_*ci*_ ∼ Bernoulli(*p*_*ci*_)

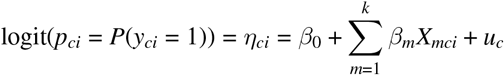

where:

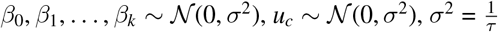,and *τ* ∼ Gamma(0.001, 0.001).

The MCMC simulation is conducted using 25,500 iterations with 500 initial burnings, 3 chains and a single thinning interval.

### 2.6 Performance metrics: Bias, Variance and Mean Square Error (MSE)

To compare the performance of the delta method, bootstrap, and MCMC approaches for estimating the variance of the Log-SIR, we evaluated several criteria such as bias (the difference between the expected value of the estimator and the true value), consistency (the estimator should converge to the true value as the sample size increases), and MSE (for overall accuracy).

## 3 RESULTS

### 3.1 Variance of Log-SIR using the three estimation methods

Summary of bias along with variance and mean square error (mse) are shown in Table 1.

**TABLE 1.** Comparison of Bias, Variance, and MSE for Different Estimation Methods.

| Method | Bias | Variance | MSE |
| --- | --- | --- | --- |
| Delta | 0.01927454 | 1.696437e-04 | 0.0005411516 |
| Bootstrap | 0.02566281 | 2.771867e-04 | 0.0009357665 |
| Bayesian | 0.01922758 | 5.142122e-05 | 0.0004211210 |

The analysis result indicated that the bias values across all methods to be more or less similar with MCMC slightly showing the lowest bias (0.01922), followed by the delta-method (0.01927) and the Bootstrap (0.02567), respectively. This suggests that the Bayesian MCMC method provides a slightly less biased variance estimate of Log-SIR than the other methods. In addition, the Bayesian MCMC exhibits the lowest variance (0.00005), indicating more stable and less dispersed estimates of the Log-SIR. Higher variance was observed in the Delta-method (0.00016), while the Bootstrapping approach resulted in the highest variance (0.00027) introducing more uncertainty in the Log-SIR estimates. Looking at the overall accuracy of the methods, the Bayesian MCMC had the lowest MSE (0.00042), indicating better overall accuracy. The delta-method follows with an MSE of 0.00054, and the Bootstrap had the highest MSE (0.00094), showing it to be the least reliable method among the methods compared.

The result, in general indicated lower values on bias, variance and MSE values. Lower bias values indicated that the estimators are more accurate on average, lower variance indicated that the estimators to be more consistent and lower MSE indicated that the estimator are both accurate and consistent. However, the parameter estimates were the lowest for the MCMC method indicating that the Bayesian approach to be more preferred approach for the estimation of the variance of the Log-SIR; var(Log-SIR). MCMC is the best-performing method as it has the lowest bias, variance, and MSE. The delta Method performs reasonably well but has slightly higher variance and MSE than MCMC. Bootstrap captures variability well but introduces more uncertainty, as seen in its high variance and MSE.

In addition, a comparison of the three methods in terms of consistency is shown in Figure 1. Accordingly, Figure 1 highlights the trade-offs among the variance estimation methods. While bootstrapping tends to be more variable, MCMC provides more stable estimates, and the Delta Method offers computational efficiency but can be less precise. Bootstrapping (Green) shows higher variance. The green points, representing bootstrap-based variance estimates, are often higher compared to the other two methods. This suggests that bootstrapping introduces additional variability, which is expected since it resamples the data and can exaggerate variance in small samples.

**FIGURE 1.**
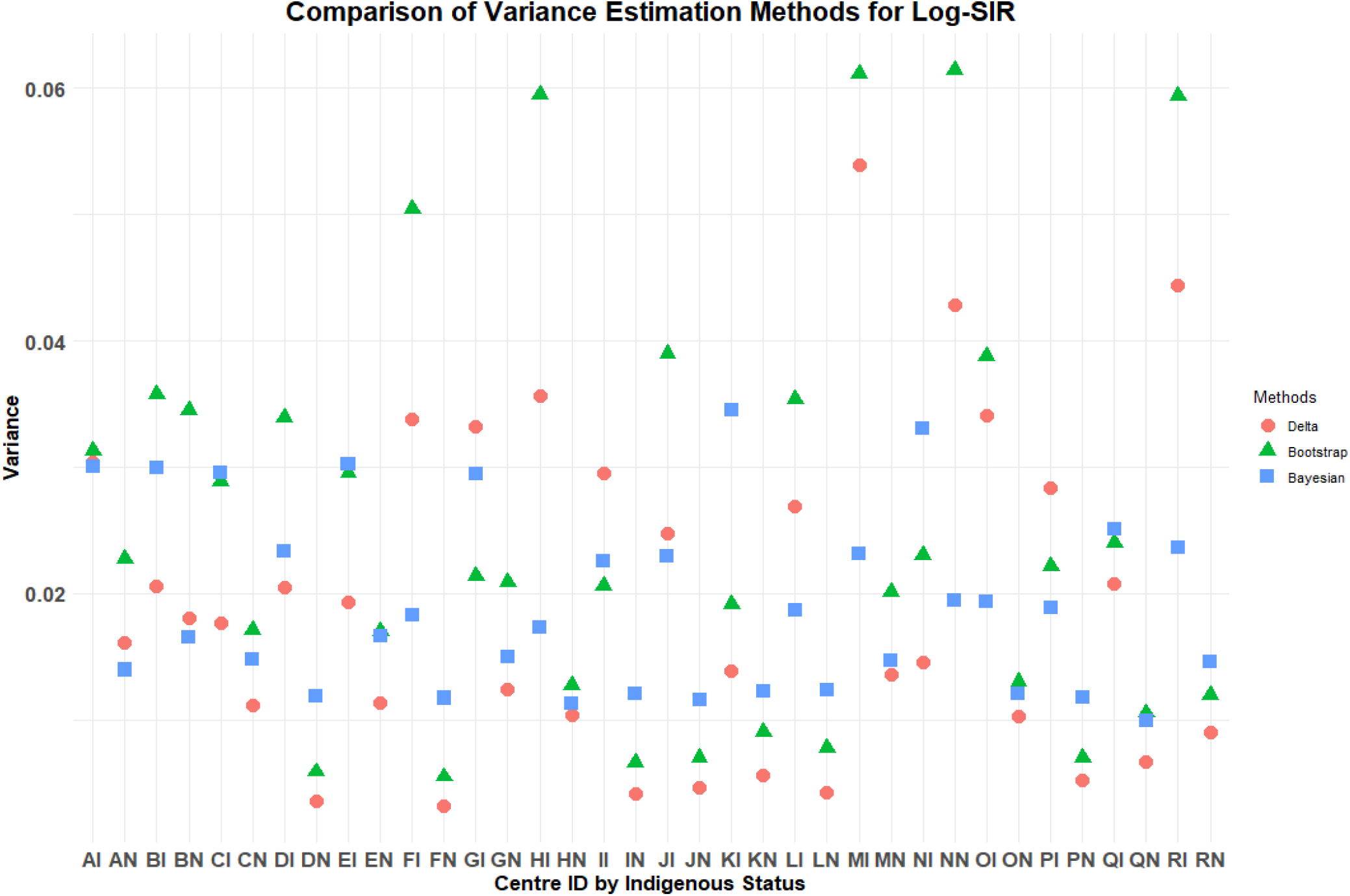
Delta-method, Bootstrapping and Bayesian approaches.

However, the Bayesian MCMC estimates (the Blue points) are more stable. They are generally lower than bootstrapping but slightly higher than the Delta-method in for most of the cases. The Bayesian methods incorporate prior information, and this leads to more stabilized variance estimates.

Delta-method (Red) is the most conservative and hence it often yields the lowest variance estimates. This method uses first-order approximations and may underestimate variance, especially for complex or skewed data distributions.

A summary table for each centre is shown in Table 2. As it is evident from Table 2, the standard errors are high variable across centres using the bootstrap method followed by the delta-method.

**TABLE 2.** Standard error of Log-SIR by centres using the three methods: ‘Return to Country’ outcome.

| Centre | Standard Error |  |  |
| --- | --- | --- | --- |
|  | Delta Method | Bootstrap | Bayesian |
| AI | 0.03034 | 0.03129 | 0.03003 |
| AN | 0.01609 | 0.02272 | 0.01395 |
| BI | 0.02053 | 0.03573 | 0.02997 |
| BN | 0.01805 | 0.03453 | 0.01655 |
| CI | 0.01761 | 0.02887 | 0.02955 |
| CN | 0.01116 | 0.01710 | 0.01474 |
| DI | 0.02044 | 0.03393 | 0.02329 |
| DN | 0.00356 | 0.00588 | 0.01190 |
| EI | 0.01930 | 0.02958 | 0.03021 |
| EN | 0.01129 | 0.01699 | 0.01659 |
| FI | 0.03381 | 0.05042 | 0.01832 |
| FN | 0.00318 | 0.00548 | 0.01172 |
| GI | 0.03318 | 0.02135 | 0.02946 |
| GN | 0.01242 | 0.02085 | 0.01494 |
| HI | 0.03566 | 0.05951 | 0.01728 |
| HN | 0.01038 | 0.01275 | 0.01132 |
| II | 0.02950 | 0.02063 | 0.02259 |
| IN | 0.00416 | 0.00657 | 0.01205 |
| JI | 0.02471 | 0.03898 | 0.02294 |
| JN | 0.00459 | 0.00698 | 0.01157 |
| KI | 0.01382 | 0.01916 | 0.03451 |
| KN | 0.00555 | 0.00901 | 0.01221 |
| LI | 0.02685 | 0.03541 | 0.01869 |
| LN | 0.00423 | 0.00774 | 0.01231 |
| MI | 0.05387 | 0.06111 | 0.02316 |
| MN | 0.01353 | 0.02013 | 0.01468 |
| NI | 0.01449 | 0.02302 | 0.03302 |
| NN | 0.04283 | 0.06143 | 0.01941 |
| OI | 0.03405 | 0.03878 | 0.01934 |
| ON | 0.01028 | 0.01302 | 0.01206 |
| PI | 0.02837 | 0.02216 | 0.01889 |
| PN | 0.00521 | 0.00696 | 0.01179 |
| QI | 0.02078 | 0.02396 | 0.02511 |
| QN | 0.00669 | 0.01053 | 0.00990 |
| RI | 0.04436 | 0.05934 | 0.02358 |
| RN | 0.00900 | 0.01197 | 0.01455 |

In summary, there are notable variations in variance estimates across centres. Some centres exhibit more spread between methods, suggesting that the choice of method affects variance estimates significantly.

In the next section, we presented funnel plots constructed using the three methods for assessing centres’ performance in providing services close to home for patients with ESKD. The main focus is to highlight how the variance estimation methods provide somehow variable plots and how they affect interpretation and decision making on the performance of centres in service provision.

### 3.2 Centres’ performance using funnel plots

A summary funnel plot using the three methods is displayed in Figure 2, 3 and 4. Each funnel plot has different variance estimates for the same underlying data. The funnel plots evaluate centre-level performance in treating ESKD patients close to home by comparing the Log-SIR across different centres stratified by Indigenous status. The x-axis represents effective sample size (defined as a measure of the variability of the Log-SIRs for each centre relative to the total variability of all Log-SMRs (Kasza et al., 2013) ^38^), while the y-axis measures Log-SIR, indicating whether observed rates of receiving treatment close to home are higher or lower than expected. Centres within the upper and lower FDRs indicate expected performance in treating patients close to home (are in the region of average performance). The dashed lines forming funnels around the horizontal solid line (Log-SIR=0) indicate expected variation, with centres falling outside these limits exhibiting statistically significant differences from the norm.

**FIGURE 2.**
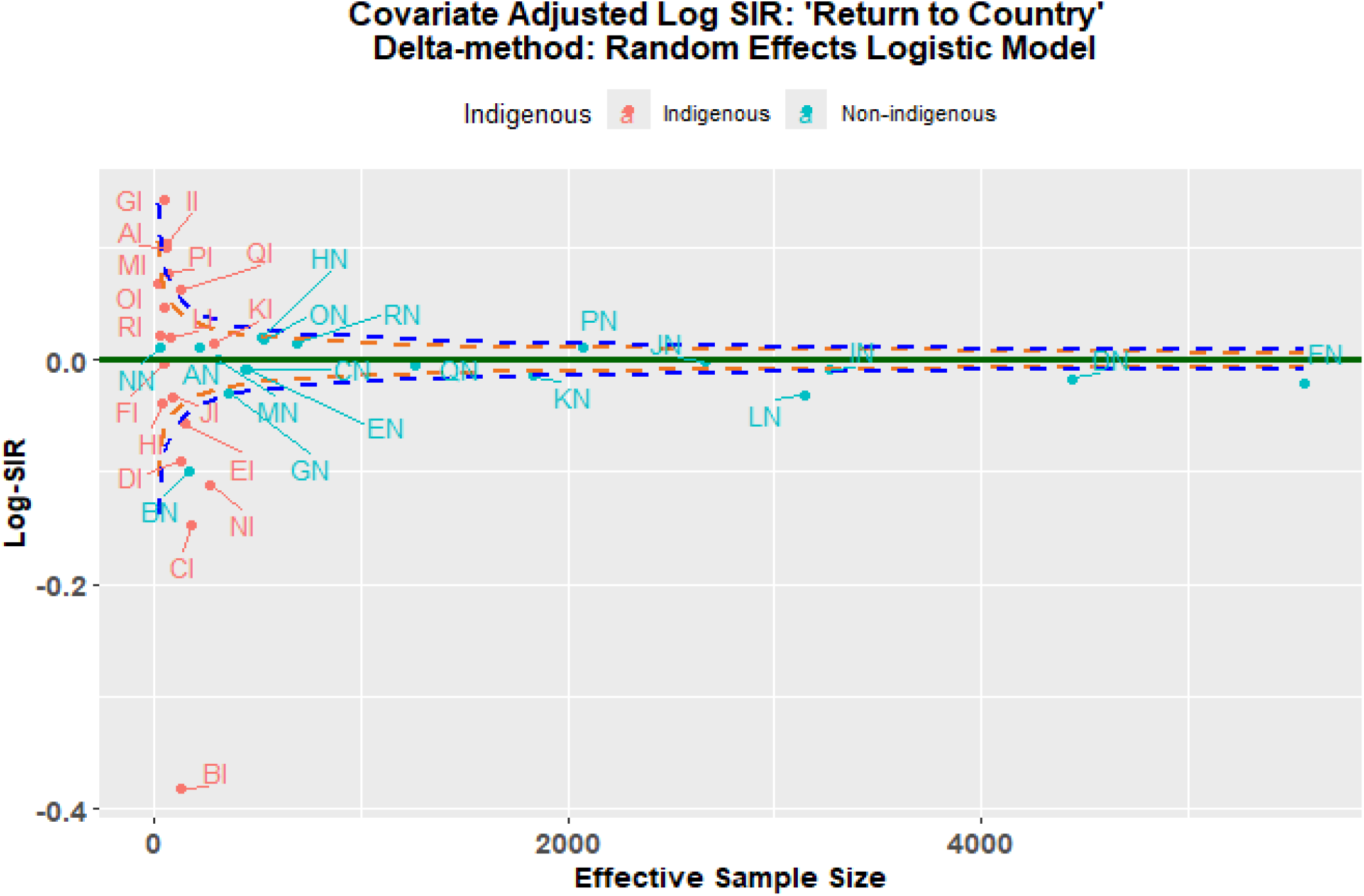
Funnel plot using Delta-method.

**FIGURE 3.**
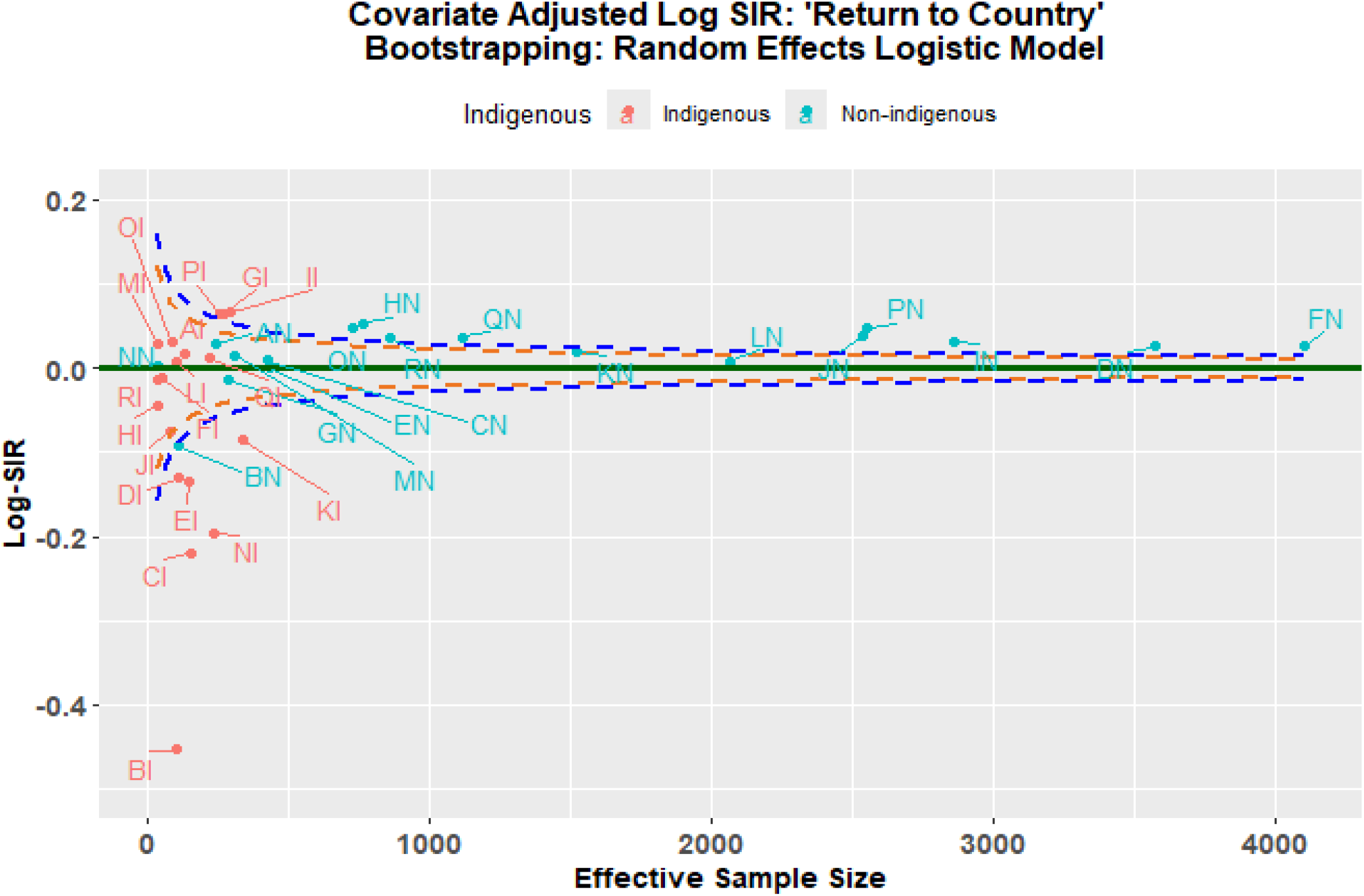
Funnel plot using Bootstrapping.

**FIGURE 4.**
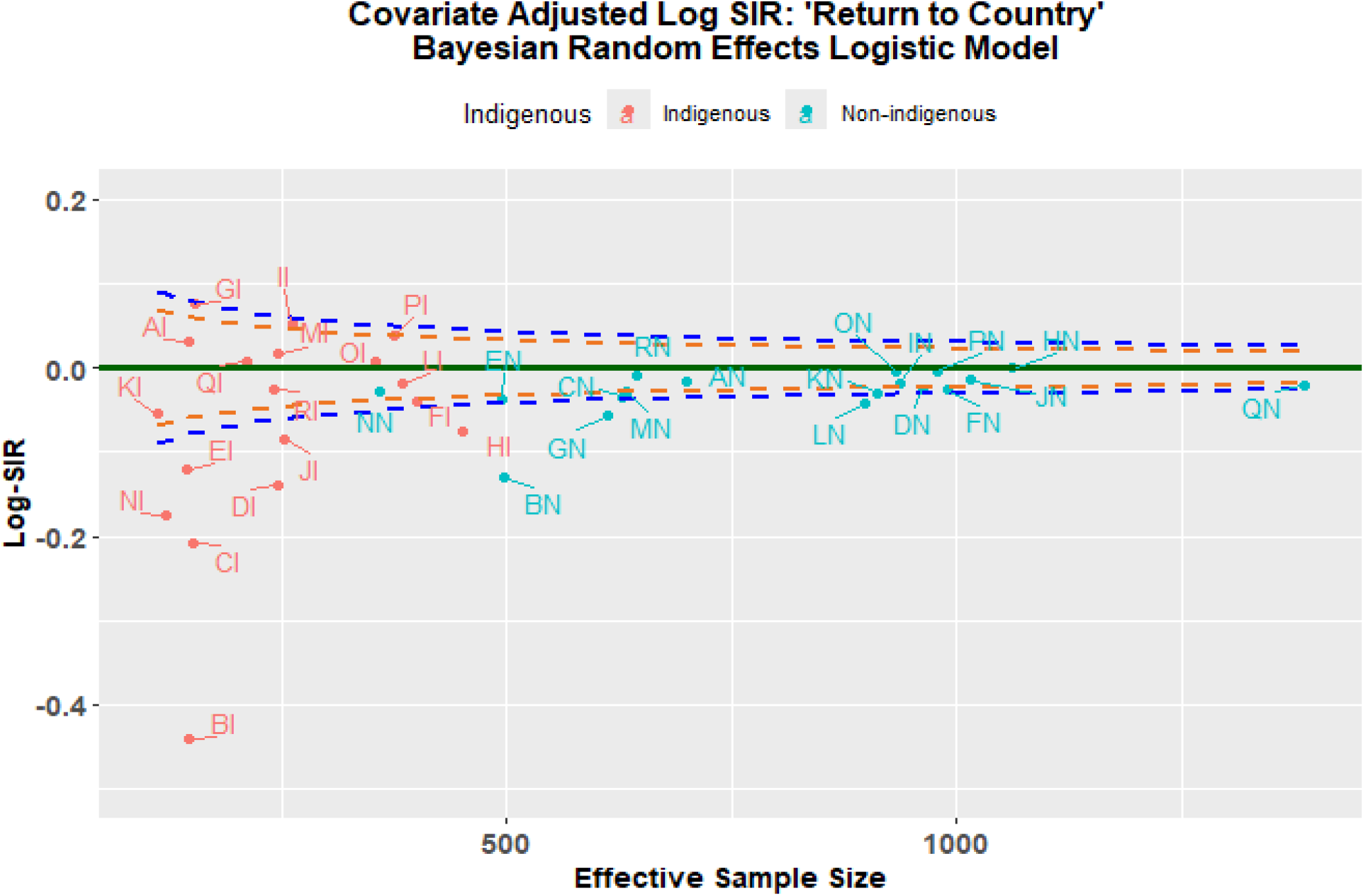
Funnel plot using Bayesian MCMC.

Figure 2 presents a funnel plot that compares the performance of centres using delta-method for estimating the variance of the Log-SIR. Centres within the upper and lower FDRs indicate expected performance in treating patients close to home (are in the region of average performance). The dashed lines forming funnels around the horizontal solid line (Log-SIR=0) indicate expected variation, with centres falling outside these limits exhibiting statistically significant differences from the norm.

Using this approach, six centres, BI, BN, CI, DI, EI and NI were low performing while three centres, GI, II and QI were higher-than-average performers. The remaining centres lie within the false discovery rates being average performers in treating patients close to home. Centre BI shows the lowest Log-SIR, suggesting exceptionally lower performance in treating patients close to home. Overall, larger centres exhibit more stable Log-SIR values, while smaller centres experience greater variation, reinforcing the importance of centre size in the assessment of centres’ performance in treating patients close to home. Using this method, the variance of Log-SIR appears relatively low, with most values concentrated around zero.Some extreme values (outliers) are present on the left-hand side, indicating a few centres with more deviation. The spread of points suggests that this method results in a tighter distribution of Log-SIR.

Figure 3 compares centres using bootstrapping approach. Using this method, seven centres, BI, BN, CI, DI, EI, NI and KI were lower-than-average performers. However, twelve centres, GI, II, PI, DN, FN, HN, IN, JN, ON, PN, QN, and RN, were found to be higher-than-average performing. The remaining centres lie within the false discovery rates being average performers in treating patients close to home. Notably, BI remains an outlier with the lowest Log-SIR, reflecting exceptionally low performance in treating patients close to home. Using bootstrap, the variance is slightly larger compared to the first plot. The spread of Log-SIR values is more noticeable, with a wider range of deviations from zero.Morecentres have larger deviations, particularly on the left side, compared to the delta-method.

Figure 4 presents a funnel plot that compares the performance of centres using Bayesian approach for estimating the variance of the Log-SIR. Accordingly, eleven centres, BI, BN, CI, DI, EI, GN, HI, JI, KI, NI, and LN were found low-than-expected performers and no centre was found to be top performing in treating patients close to home. Larger centres exhibit more stable Log-SIR values, reinforcing the reliability of their performance assessments. Using the Bayesian approach, the variance of Log-SIR is still larger than the first plot but somewhat comparable to the second. The spread is not as extreme as in the second plot, but it still shows noticeable deviations.There are clear differences in the spread of values across regions.

The delta-method results in the least variance in Log-SIR while the bootstrapping has the highest variance, with a wider spread of values. Clearly, the Bayesian approach has an intermediate variance, showing more spread than the first method but less than the second.

In summary, these results highlight centre-level differences in treating patients close to home, and this is coupled with variability in variance estimation by the three methods. The stability of the Log-SIR using the Bayesian approach could be due to the fact that Bayesian method borrow strength from prior beliefs summarised using probability distributions that smooth variability in estimation.

## 4 DISCUSSION

In healthcare providers’ performance assessment, standardized incidence ratios (SIRs) and standardized mortality ratios (SMRs) are essential tools used to assess whether observed rates of disease or death deviate from what is expected. Accurate estimation of variance in these ratios is crucial as it affects decision-making regarding providers’ performance, resource allocation, and quality improvement strategies. In this paper, we compared three methods, namely, delta-method, bootstrapping and Bayesian approach, to estimate the variance of the Log-SIR given by equation (3) and considered funnel plot approaches to build false discovery rates around the Log-SIR using these three variance estimators. The variance estimation methods have been widely discussed in statistical literature. Gelman et al. ^33^ emphasize that Bayesian methods, particularly MCMC, provide more stable estimates due to their ability to incorporate prior information and reduce uncertainty. Similarly, Efron and Tibshirani ^11^ discuss bootstrapping as a flexible but sometimes overly variable approach, which aligns with our findings of increased variance in bootstrapped estimates. The delta-method is frequently used in epidemiology for variance estimation Rothman, et al ^34^ . It provides an efficient and straightforward way of estimating the variance of Log-SIR or Log-SMR, especially when the distribution of the underlying data correctly specified. This method can be computationally efficient, but its accuracy may suffer in cases where the underlying distribution deviates significantly from the assumed form Canan, et al. ^35^ . When applied in healthcare decision-making, such as assessing the performance of hospitals based on SMRs, the delta-method may underestimate variance if assumptions are violated. This could lead to incorrect conclusions regarding the performance of healthcare providers.

Variance estimation using the delta-method for metrics other than SMR have used intensively. For instance, Sharon-Lise & David ^32^ applied the delta-method to approximate the variance of demographic parameters in avian biology studies. Although not directly related to healthcare, this study illustrates the broader applicability of the delta-method in estimating variances of complex ratios. Also, Lee, et al (2024) compared Green, delta-method and monte carlo methods for calculating the 95% confidence interval for population-attributable fraction (PAF) ^36^. In addition, Sauer, et al. (2020) applied delta-method for variance estimation for effective coverage measures ^37^. There is limited study that directly applied delta-method in the estimation of Log-SIR used in the assessing performance of health care providers in the provision of health services for a given outcome. Bootstrapping, on the other hand, has the advantage of not relying on distributional assumptions and can be used to directly estimate the distribution of Log-SIR or Log-SMR. This can lead to more robust variance estimates, particularly in settings with small sample sizes or unknown distributions. By resampling, bootstrapping accounts for sampling variability and can help improve the precision of performance assessments ^11^. For instance, Kasza, et al. (2013) used bootstrapping for evaluating the performance of Australian and New Zealand intensive care units in 2009 and 2010 quantified by the standardised mortality ratio ^38^. Moreover, Walters, et al. (2004) used bootstrap methods for analysing health-related quality of life outcomes used in clinical trials as primary outcome measures. They found that certain bootstrap methods provided more accurate variance estimates, especially when the distribution of the outcome is unknown or ordinal scale ^39^.

In contrast, Bayesian methods provide a full posterior distribution for variance estimates, allowing for the incorporation of prior knowledge, such as expert opinion or historical data on hospital performance. This can lead to more flexible and informative variance estimation, especially when data are sparse or prior knowledge is available. Bayesian methods can also be used to model hierarchical structures (e.g., hospitals within regions), providing more precise estimates of performance at various levels ^28^.

A study by George, et al. ^40^ applied Bayesian hierarchical models to estimate hospital performance in the Hospital Compare model for acute myocardial infarction (AMI) mortality. They found that indirect standardization fails to adequately control for differences in patient risk factors and systematically underestimates mortality rates at the low volume hospitals.

Our results showed that Bayesian approaches provided more conservative estimates with tighter credible intervals, particularly in hospitals with small case volumes. We demonstrated that Bayesian MCMC outperforms the other methods in terms of lower variance and MSE, making it the preferred choice for estimating Log-SIR variance when computational resources permit.

Below we have summariesed the variability in variance estimates and their implications on funnel plots and epidemiological studies.

### 4.1 Variability in log SIR Variance Estimates

The three methods yield different variance estimates for the same underlying data. Bootstrapping tends to produce higher variance estimates due to the nature of resampling, which can exaggerate variability, particularly in small samples ^41^. In contrast, Bayesian (MCMC) estimates tend to be more stable, benefiting from prior distributions that help regularize estimates, a characteristic also observed in Bayesian hierarchical models for disease mapping ^42^. The delta-method, being a first-order approximation, is the most conservative, often producing the lowest variance estimates, which may lead to underestimation in complex data structures ^28^. These differences highlight the importance of choosing an estimation method suited to the underlying data characteristics and sample size.

In our study, the variance estimates differ across methods, with bootstrapping tending to show more extreme values (both high and low) compared to the other two methods. MCMC appears to provide more stable and generally lower variance estimates compared to bootstrapping. Delta method is relatively consistent but tends to lie between the MCMC and bootstrap estimates. Some centres have noticeably higher variance estimates for all three methods (e.g., locations where green dots are well above the others). This suggests that uncertainty in Log-SIR estimation varies by centre, possibly due to differences in sample size, population characteristics, or underlying risk factors. Bootstrapping shows more variability, which is expected since it resamples data and may amplify variability in small samples. MCMC provides more stable estimates, benefiting from Bayesian shrinkage and prior information incorporation. The delta method is computationally efficient but may underestimate variance in some cases (e.g., when normality assumptions are violated) ^43^. Centres with higher variance estimates (especially under bootstrapping) suggest that Log-SIR estimates are more uncertain there, which should be considered when making public health decisions. If variance estimates are too high, it may indicate the need for larger sample sizes or improved data collection in those centres.

### 4.2 Impact on Funnel Plots

The funnel plots illustrate how these methods influence the distribution of Log-SIR estimates. The Bayesian approach exhibits a more stabilized pattern, particularly at smaller sample sizes, where shrinkage effects help reduce extreme values. This aligns with findings from Spiegelhalter et al. ^44^, who demonstrated that Bayesian hierarchical modelling effectively mitigates overdispersion in epidemiological data. Conversely, the bootstrapping approach results in greater spread at smaller sample sizes, reflecting its sensitivity to sample fluctuations. Similar findings have been reported in comparative studies on variance estimation methods, where bootstrapping is noted to introduce greater variability but remains valuable for robust uncertainty estimation ^11^ Although both methods show convergence of Log-SIR estimates toward zero as sample sizes increase, bootstrapping maintains slightly higher variance, reinforcing the need for careful interpretation in small-sample studies.

### 4.3 Implications for Epidemiological Studies

The choice of variance estimation method has significant implications for epidemiological research. Bayesian methods offer improved stability and are particularly useful when incorporating prior knowledge is beneficial. Studies have shown that Bayesian approaches reduce estimation bias and enhance interpretability in spatial epidemiology ^45^. Bootstrapping, despite its higher variability, remains a valuable tool for robust uncertainty estimation, especially when parametric assumptions may not hold ^46^. Meanwhile, the delta-method, though computationally simple, may underestimate variance, making it less reliable for complex data scenarios, as previously noted in statistical inference literature ^28^. These findings align with broader discussions on variance estimation in epidemiology, emphasizing the trade-offs between robustness, computational efficiency, and precision ^47^.

These findings highlight the importance of selecting an appropriate variance estimation method depending on the study context. Bayesian methods may be preferable when stability and regularization are critical, while bootstrapping is useful for assessing variability in more flexible settings. The delta-method should be employed cautiously, particularly when dealing with skewed or complex distributions. Future research should explore hybrid approaches that combine the strengths of these methods for more robust inference ^11,28^

Our study has several limitations. Firstly, while understanding the differences between variance estimation methods is crucial for assessing the reliability of standardized incidence ratio (SIR) estimates across different centres, we didn’t consider how model (Logistic vs Poisson) choice influence variance estimates and hence the resulting statistical infrence. Secondly, we only considered non-parameteric bootstrapping and implications of the parametric bootstrapping was not assessed. Third, we didn’t consider other transformations than logarithmic transformation and their effect on the interpretation of providers’ performance. For instance, Quaresma, et al ^8^ investigated the implications of identity(log), complementary log-log, logit and logarithmic transformation in their study of cancer survival. Finally, within the random effects logistic regression, we only considered logit link and other links such as probit and complementary log-log link were not considered here.

## 5 CONCLUSIONS

In conclusion, the choice of variance estimation method plays a significant role in how healthcare providers’ performance is assessed. While each method has its strengths and weaknesses, bootstrapping and Bayesian approaches generally provide more reliable estimates of uncertainty compared to the delta-method. However, the choice of method should consider computational resources, data structure, and the available prior knowledge for Bayesian methods. Decision-makers should be aware of the implications of variance estimation on conclusions regarding provider performance, which can influence policy, resource allocation, and quality improvement initiatives in healthcare settings. In terms of decision-making, the choice of variance estimation method can affect the conclusions drawn about the performance of healthcare providers. Using the delta-method may lead to an underestimation of uncertainty, especially when the data do not meet distributional assumptions. Bootstrapping, while more robust, may be computationally intensive, especially with large datasets. Bayesian methods, with their flexibility and ability to incorporate prior knowledge, can be powerful tools but require careful specification of priors and may be computationally demanding.

## Data Availability

All data produced in the present study are available upon reasonable request to the authors

## Abbreviations

SIR: Standardised Incidence Ratio
SMR: Standardised Mortality Ratio
Log-SIR: Logarithm of the Standardised Incidence Rati
SD: Standard Deviations
MSE: Mean Square Errors
MCMC: Markov Chain Monte Carlo
ANZDATA: Australia and New Zealand Dialysis and Transplant Registry
KRT: Kidney Replacement Therapy
FDR: False Discovery Rate

## AUTHOR CONTRIBUTIONS

SW and PL conceived the idea of the research, collected and analyzed the data. SW prepared the first and subsequent drafts. PL and YJ participated in data interpretation and provided comments during subsequent drafts. All authors read and approved the final manuscript.

## ACKNOWLEDGMENTS

We are grateful to the Australian National Health and Medical Research Council (NHMRC)for supporting the “Return to Country” project (GNT1158075) of which this methodological paper is a part of. The data reported here have been supplied by the Australia and New Zealand Dialysis and Transplant Registry (ANZDATA). The interpretation and reporting of these data are the responsibility of the authors and in no way should be seen as an official policy or interpretation of the Australia and New Zealand Dialysis and Transplant Registry.

## SUPPORTING INFORMATION

Additional supporting information may be found in the online version of the article at the publisher’s website.

**How to cite this article:** Woldeyohannes S., Jones Y, Lawton P. Variance Estimation for Assessing Healthcare Providers’ Performance using log Standardized Incidence Ratio. *Statistics in Medicine*. 2025;00(00):1–18.

**APPENDIX**

### A. MODEL AND LOG SIR DEFINITIONS

In the following we presented model specification, the derviation of the variance for the Log-SIR using the delta-method, and a description of the bootstrap and Bayesian approaches for estimating variance of Log-SIR.

#### A.1 Model specification

Since we have binary outcome of receiving treatment close to home for ESKD, denoted by *y*_*ci*_, from *n*_*c*_ number of patients receiving treatment from the *c*^*th*^ centre/hospital. Where, *c*, …, *N* centres, we proposed a Bernoulli sampling distribution for the probability of getting treatment close to home for the *i*^*th*^ patient from the *c*^*th*^ centre.

That is, *y*_*ci*_ ∼ Bernoulli(*p*_*ci*_) and a random effects logistic regression model can be specified as:

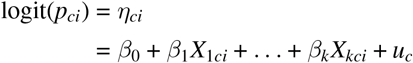

where:

- *y*_*ci*_ is the binary outcome for patient *i* in centre *c*.
- *X*_1*ij*_ … *X*_*kij*_ are *k* covariates for patient *i* in centre *c*.
- *β*_0_, *β*_1_, …, *β*_*k*_ are fixed effects.
- *u*_*c*_ is the random effect for centre *c*, assumed to be normally distributed: *u*_*c*_ ∼ 𝒩 (0, *σ*^2^).

In a random effects model, the random effects *u*_*c*_ are assumed to follow a normal distribution:

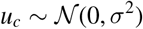

The density function for the normal distribution is:

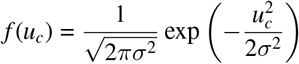

The probability of observing *y*_*ci*_ given the fixed and random effects is:

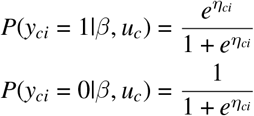

where *η*_*ci*_ = *β*_0_ + *β*_1_*X*_1*ci*_ + … + *β*_*k*_*X*_*kci*_ + *u*_*c*_ is the linear predictor. The probability for a single patient *i* in centre *c* is:

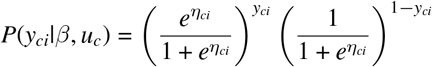

The fixed effects *β* and the variance parameter 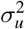 can be estimated using maximum likelihood estimation (MLE) through the ‘glmer’ function in R.

#### A.2 Standardized Incidence Ratio (SIR)

In the next, we presented details of the derivation of the final formula for the variance of log(SIR) using the delta method given the sampling distribution and the model formulations above.

The SIR for the *c*^*th*^ centre is defined as:

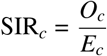

where,

- *O*_*c*_: The sum of the observed outcomes (receving treatment close to home) for all patients in centre *c*:

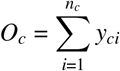

where *y*_*ci*_ is the binary outcome (1 if the patient is receiving treatment at home, 0 otherwise) for patient *i* in centre *c*.
- *E*_*c*_: The sum of the predicted probabilities of receving treatment close to home for all patients in centre *c*:

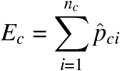

where 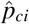 is the predicted probability of receving treatment close to home for patient *i* in centre *c*.

The Log-SIR_*c*_ is given by:

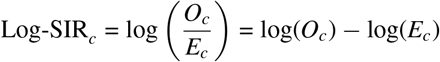

### B. DELTA-METHOD

The delta method is a technique used to approximate the variance of a function of one or more random variables. In the context of estimating the variance of the log standardized incidence ratio Log-SIR, we can apply the delta method to approximate the variance of log 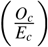.It approximates the variance of a function of random variables by using the Jacobian matrix and the covariance matrix of the original variables. For our case, the variance of Log-SIR_*c*_) is approximated by:

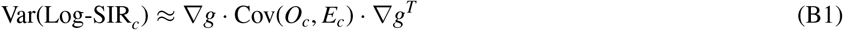

First, let’s define the covariance matrix of *O*_*c*_ and *E*_*c*_:

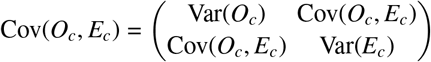

The Jacobian matrix (gradient) ∇*g* of the function 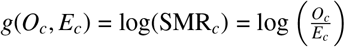 with respect to *O*_*c*_ and *E*_*c*_ is derived as:

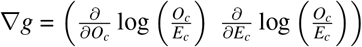

And calculating the partial derivatives with respect to the quantities, we get:

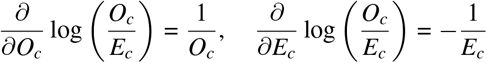

Therefore, the Jacobian matrix is:

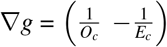

Substituting ∇*g* and Cov(*O*_*c*_, *E*_*c*_) into formula (9), we get:

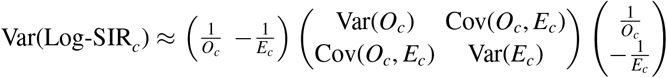

To simplify the derivation, we perform the matrix multiplication step by step: First, multiply the Jacobian matrix by the covariance matrix:

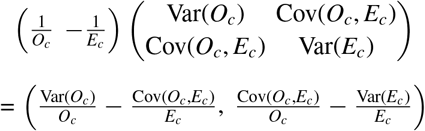

Then, multiply the resulting vector by the transpose of the Jacobian matrix:

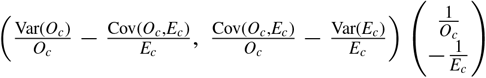

Finally we, get the Var(Log-SIR_*c*_) shown in equation (3)

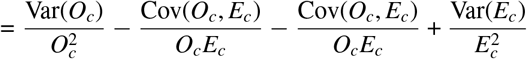

In the next, we will derive estimates for the Var(O_*c*_), Var(E_*c*_) and Cov(O_*c*_, E_*c*_).

#### B.1 Variance of *O*_*c*_: Var(O_*c*_)

The observed count *O*_*c*_ for a centrel *c* is defined as the sum of individual binary outcomes *Y*_*i*_:

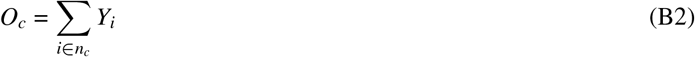

where *Y*_*i*_ is a Bernoulli random variable (taking values 0 or 1), indicating whether an event occurs for individual *i*.

Since *O*_*c*_ is the sum of correlated random variables, its variance expands as:

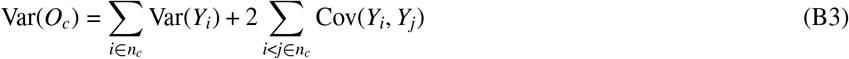

This follows from the general variance formula for sums of correlated random variables:

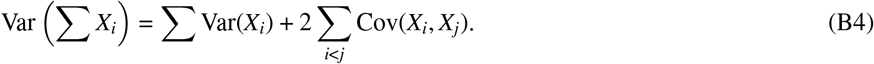

##### Variance of *Y*_*i*_

For a binary outcome *Y*_*i*_ with probability *p*_*i*_ = *P*(*Y*_*i*_ = 1), the variance is:

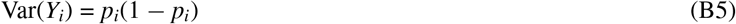

which is the standard variance formula for a Bernoulli-distributed variable.

##### Covariance Between *Y*_*i*_ and *Y*_*j*_

To derive the covariance term Cov(*Y*_*i*_, *Y*_*j*_), we assume a logistic regression model with random intercepts. The probability *p*_*i*_ is modeled as:

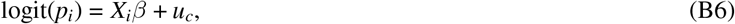

where:

- *X*_*i*_ are fixed covariates,
- *β* is the fixed effect coefficient,
- *u*_*c*_ is the random effect for centre *c*, assumed to be normally distributed as 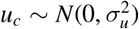.

Since *Y*_*i*_ and *Y*_*j*_ share the same random effect *u*_*c*_, they become correlated through *u*_*c*_.

Using standard results from generalized linear mixed models (GLMMs), the covariance between two binary outcomes *Y*_*i*_ and *Y*_*j*_ in the same group (centre *c*) is approximately:

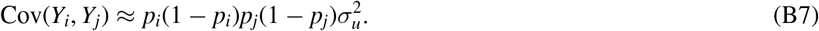

This approximation comes from the fact that when a common random effect *u*_*c*_ influences both *Y*_*i*_ and *Y*_*j*_, their correlation is roughly proportional to the variance of the random effect, 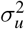.Specifically:

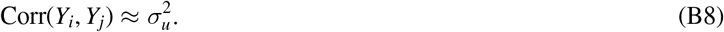

Multiplying by the standard deviations of *Y*_*i*_ and 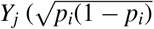 and 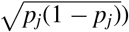,we obtain:

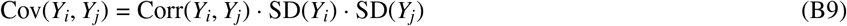

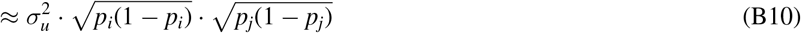

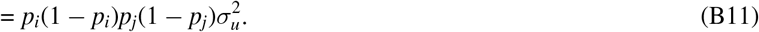

##### Final Expression for Variance of *O*_*c*_

Substituting the variance and covariance terms into the variance expansion formula, we obtain:

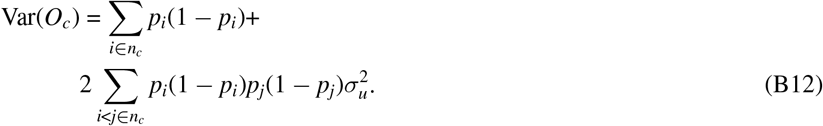

This accounts for both the independent variation in individual outcomes and the correlation induced by the shared random effect.

#### B.2 Variance of *E*_*c*_: Var(E_*c*_)

The expected counts *E* are the sum of predicted probabilities *p*_*i*_ for individuals within a hospital. The variance of *E* arises from the uncertainty in the predicted probabilities due to the random effects.

##### Derivation of Var(*E*_*c*_)

The expected counts for centrer *c* are:

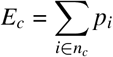

The variance of *E*_*c*_ is:

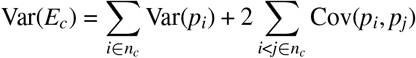

For the random-effects logistic regression model:

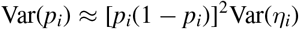

where 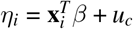 is the linear predictor. The covariance between *p*_*i*_ and *p*_*j*_ (for *i ≠ j*) is:

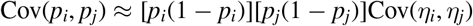

Since *η*_*i*_ and *η*_*j*_ share the same random effect *u*_*c*_:

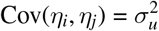

Thus:

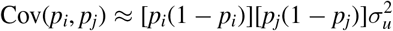

Combining these results:

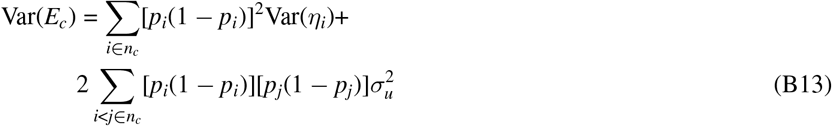

#### B.3 Covariance of *O*_*c*_ and *E*_*c*_**: Cov(O**_*i*_, **E**_*i*_)

The covariance between *O* and *E* arises because both depend on the same underlying probabilities *p*_*i*_, which are influenced by centre level shared random effect.

##### Derivation of Cov(*O*_*c*_, *E*_*c*_)

To derive the covariance Cov(*O*_*c*_, *E*_*c*_), where *O*_*c*_ is the observed count and *E*_*c*_ is the expected count, we have the definitions:

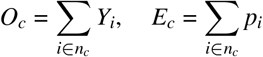

where *Y*_*i*_ is a binary outcome indicating an event occurrence for individual *i* in centre *c*, and *p*_*i*_ is the corresponding probability.

Using the linearity property of covariance, the covariance between *O*_*c*_ and *E*_*c*_ is:

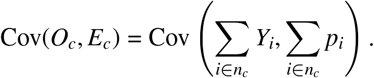

And if we expand using the property of covariance for sums, we get:

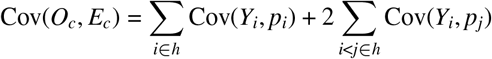

In the following, we will derive Cov(*Y*_*i*_, *p*_*i*_) and Cov(*Y*_*i*_, *p*_*j*_) for *i ≠ j*

##### Cov(*Y*_*i*_, *p*_*i*_)

Since *Y*_*i*_ follows a Bernoulli distribution:

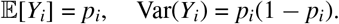

The probability *p*_*i*_ depends on the linear predictor *η*_*i*_, which includes both fixed and random effects:

*η*_*i*_ = *X*_*i*_*β* + *u*_*c*_,

where *X*_*i*_ represents the fixed effect covariates and *u*_*c*_ is the hospital-specific random effect.

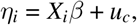

Since *p*_*i*_ = logit^−1^(*η*_*i*_), a first-order Taylor series approximation around 𝔼 [*η*_*i*_] gives:

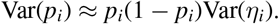

Thus,

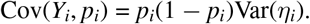

##### Cov(*Y*_*i*_, *p*_*j*_) **for** *i ≠ j*

For individuals *i* and *j* in the same hospital, their probabilities share the same random effect *u*_*c*_, inducing correlation:

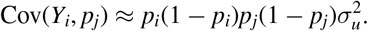

Substituting these results into our earlier equation, we obtain final expression for the Cov(*O*_*c*_, *E*_*c*_):

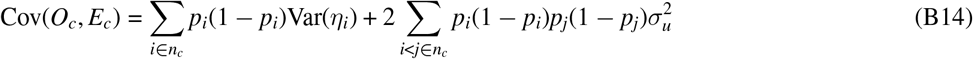

### C BAYESIAN APPROACH

Model Specification

Consider *y*_*ci*_ as a binary outcome for individual patient *i* in centre (or hospital) *c*. In addition, let *u*_*c*_ be the random effect for centre *c*. The model assumes that the probability of *y*_*ci*_ = 1 is given by a logistic regression:

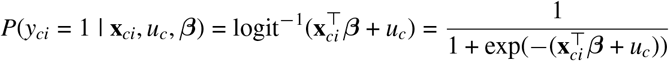

where: - **x**_*ci*_ is a vector of covariates for individual patient *i* in centre *c*. - ***β*** is a vector of fixed effects.

-The hierarchical model assumes that the random effects *u*_*c*_ are normally distributed:

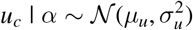

where *µ*_*u*_ and 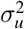 are hyperparameters (e.g., *µ*_*u*_ = 0 is common) and hence, the density function for *u*_*c*_ is the normal distribution given by:

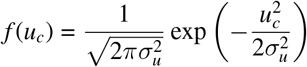

Likelihood Function

The likelihood for the binary outcome *y*_*ci*_ given the parameters ***β*** and *u*_*c*_ is:

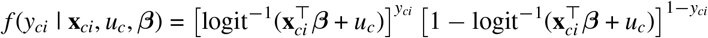

The likelihood for all observations within centre *c* is:

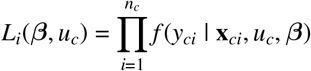

where *n*_*c*_ is the number of observations in centre *c*.

Prior Distributions

- Let’s assume that ***β*** follows a multivariate normal prior distribution:

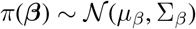

This implies that the prior density for ***β*** is:

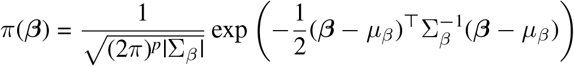

where: - *µ*_*β*_ is the mean vector for ***β***, - Σ_*β*_ is the covariance matrix for ***β***, - *p* is the dimension of ***β***.

- For the random effects, we have:

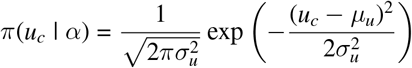
- Hyperparameters *α* = (*µ*_*u*_, *σ*^2^) might also have their own priors, but for simplicity, we’ll focus on *u*_*c*_.

Posterior Distribution

The posterior distribution for the parameters ***β*** and *u*_*c*_ given the data **y** and covariates **X** is:

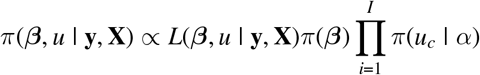

Substituting the likelihood and the prior distributions:

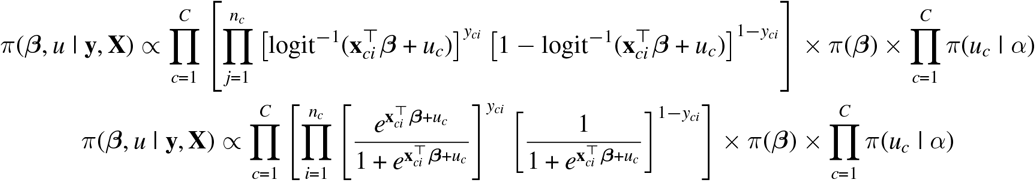

Final Posterior Distribution with Priors for ***β*** and *u*_*c*_

Now, we incorporate the prior *π*(*u*_*c*_ | *α*) into the posterior distribution:

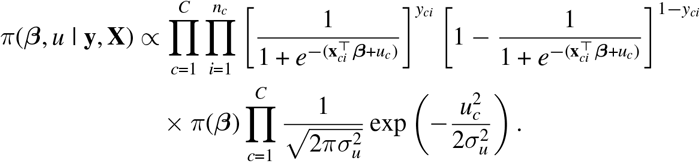

This can be written as:

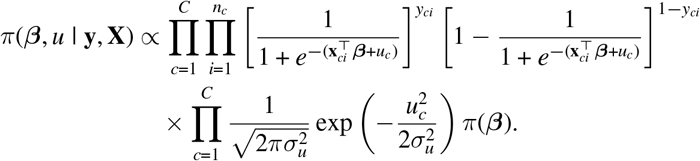

The full posterior distribution now becomes:

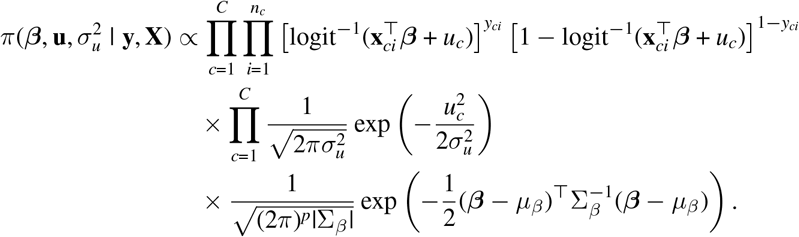

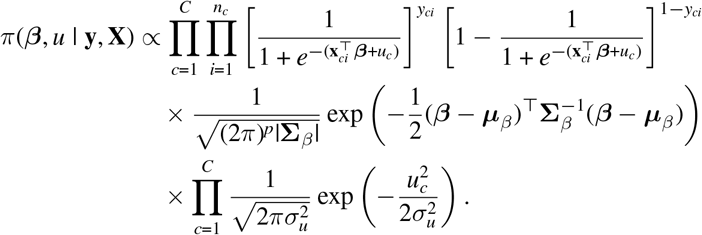

